# Solid-state esophageal pressure sensor for the estimation of pleural pressure: a bench and first-in-human validation study

**DOI:** 10.1101/2024.10.01.24314687

**Authors:** Julien P. van Oosten, Nico Goedendorp, Amne Mousa, Rutger Flink, Rik Schaart, Merel Flinsenberg, Peter Somhorst, Diederik A.M.P.J. Gommers, Leo Heunks, Annemijn H. Jonkman

## Abstract

**Background:** Advanced respiratory monitoring through the measurement of esophageal pressure (Pes) as a surrogate of pleural pressure helps guiding mechanical ventilation in ICU patients. Pes measurement with an esophageal balloon catheter, the current clinical reference standard, needs complex calibrations and a multitude of factors influence its reliability. Solid-state pressure sensors might be able to overcome these limitations.

**Objectives:** To evaluate the accuracy of a new solid-state Pes transducer (Pes_solid_). We hypothesized that measurements are non-inferior to those obtained with a properly calibrated balloon catheter (Pes_bal_).

**Methods:** Absolute and relative solid-state sensor Pes measurements were compared to a reference pressure in a 5-day bench setup, and to simultaneously placed balloon catheters in 15 spontaneously breathing healthy volunteers and in 16 mechanically ventilated ICU patients. Bland-Altman analysis was performed with nonparametric bootstrapping to estimate bias and upper and lower limits of agreement (LoA).

**Results:** Bench study: Solid-state pressure transducers had a positive bias (P_solid_ – P_ref_) of around 1 cmH_2_O for the absolute minimal and maximum pressures, and no bias for pressure swings. Healthy volunteers: the solid-state transducer revealed a bias (Pes_solid_–Pes_bal_) [upper LoA; lower LoA] of 1.58 [8.19; −5.03], −2.37 [3.96; −8.69] and 3.94 [11.09; −3.20] cmH_2_O for end-expiratory, end-inspiratory and ΔPes values, respectively. ICU patients: the solid-state transducer showed a bias (Pes_solid_–Pes_bal_) [upper LoA; lower LoA] during controlled / assisted ventilation of: −0.15 [1.39; −1.70] / −0.20 [5.02; −5.41], 0.32 [3.35; −2.72] / −0.54 [4.60; −5.68] and 0.47 [3.79; −2.85] / 0.35 [3.88; −3.18] cmH_2_O for end-expiratory, end-inspiratory and ΔPes values, respectively. LoA were <2cmH_2_O for static measurements on controlled ventilation.

**Conclusions:** The novel solid-state pressure transducer showed good accuracy on the bench, in healthy volunteers and in ventilated ICU-patients. This could contribute to the implementation of Pes as advanced respiratory monitoring technique.

**Trial registration:** Clinicaltrials.gov identifier: NCT05817968 (patient study). Registered on 18 April 2023.

## INTRODUCTION

Advanced respiratory monitoring through the measurement of esophageal pressure (Pes) as a surrogate for pleural pressure helps understanding partitioned respiratory mechanics and breathing effort in mechanically ventilated patients, and could guide the individualization of ventilator settings.[1] Despite the recognized benefits, widespread implementation of esophageal manometry is still in its infancy.[1–4] It requires (technical) expertise and the validation and calibration of balloon catheters is often challenging. For instance, optimal filling volumes vary according to the balloon type, patient factors and ventilator settings, and both excessive and insufficient balloon filling volumes dampen Pes amplitudes.[1,5] In addition, the balloon may empty over time, resulting in an underestimation of pressures. Signal dampening could also occur if compliant tubing is used to connect the catheter to the extracorporeal pressure sensor. Therefore, for correct interpretation of respiratory physiology and optimal patient/ventilator management, adequate balloon position and filling volume should be regularly confirmed with an occlusion test and adjusted accordingly.[1]

Pes catheters using a solid-state pressure transducer might be able to overcome some of the above limitations. These sensors measure Pes directly inside the esophagus, allowing a faster frequency response while not being subjected to signal dampening. Previous older studies have used such transducers, but showed unacceptable signal drifting.[6,7] These studies, however, used pressure transducers that were not correctly (temperature) calibrated. Here, we evaluate the accuracy of a new solid-state Pes catheter with a transducer that allows for both temperature and ambient pressure calibration. We hypothesized that measurements are non-inferior as compared to a correctly calibrated balloon catheter. We tested this hypothesis in a bench setup, in healthy volunteers, and in mechanically ventilated patients during controlled and assisted ventilation.

## METHODS

For additional details, see Additional file 1.

### Study design and subjects

This study consisted of: 1) a bench study (September 2023) at the manufacturer location (Pulmotech B.V., Leek, the Netherlands), 2) a prospective study in spontaneously breathing healthy volunteers (August–October 2021) at the intensive care unit (ICU) of the Amsterdam University Medical Center, Amsterdam, the Netherlands (ethics approval number METC 2020.470), and 3) a prospective study in mechanically ventilated ICU patients (October 2023 to March 2024) at the ICU of the Erasmus Medical Center, Rotterdam, The Netherlands (ethics approval number MEC-2023-0119; ClinicalTrials.gov: NCT05817968). Extensive technical development tests were performed prior to the healthy volunteer study; these data are not part of this manuscript. Written informed consent was obtained from healthy subjects and patients according to local regulations.

### Healthy volunteers

We recruited healthy, non-obese adults without history of cardiac and/or pulmonary disease and contraindications for nasogastric catheter placement (e.g., esophageal varices, recent (<2 weeks) nasal bleeding, use of anticoagulants).

### Patients

Elective adult cardiothoracic surgery patients requiring postoperative invasive controlled mechanical ventilation in the ICU were enrolled pre-surgery. Eligibility was reassessed at ICU arrival. Exclusion criteria were: (1) upper airway/esophageal/mouth or face pathology (i.e. recent surgery, esophageal varices, diaphragmatic hernia), (2) nasal bleeding within the last 2 weeks, (3) presence of pneumothorax, (4) inadequate coagulation, (5) pregnancy.

### Data collection

We collected sex, age, height, body mass index (BMI), and for the ICU population also the relevant medical history (cardiac and pulmonary diseases), type of surgery performed, vital signs and ventilator settings throughout study procedures. Device-related adverse events were noted.

### Esophageal manometry

We tested the intelligent Esophageal Pressure Catheter (iEPC) (PulmoTech B.V.), a CE-marked 12 French catheter with a length of 125cm that combines nasogastric feeding with Pes measurements via a solid-state pressure sensor (Additional file 2). The catheter was connected to an acquisition system (iEPMS, PulmoTech B.V. connected to Polybench, Applied Biosignals GmbH, Germany) for data sampling at 200Hz.

Measurements were compared with a standard balloon catheter, either the Cooper catheter (Cooper Surgical, USA: 5 French, length 85cm, balloon length 9.5cm – used in healthy volunteers and patients) or the NutriVent^TM^ catheter (Sidam, Italy: 14 French catheter, length 108cm, balloon length 10cm – used in patients). The balloon catheter, an airway pressure (Paw, healthy subjects + patients) and flow sensor (healthy subjects) were connected to an acquisition system (MP160, BIOPAC Systems Inc., USA) for simultaneous recording of waveforms sampled at 200Hz. Waveforms were synchronized with the solid-state sensor tracings offline.

### Procedures Bench

Catheters were exposed to physiological conditions (100% relative humidity and 37°C) for 5 days (Additional file 3). Pressure swings of 12 cmH_2_O above 10 cmH_2_O baseline at a rate of 12/minute were applied using the AVEA (Viasys Healthcare, USA) in conjunction with a humidifier (F&P MR850, New Zealand). Temperature and relative humidity were controlled to maintain 37±2°C and >90%, respectively. Reference pressure (P_ref_) was measured through a non-compliant tube connected to the setup. Data was recorded at 30Hz for the solid-state pressures (P_solid_), and at 200Hz for P_ref_ (iEPMS, PulmoTech B.V.).

### Healthy volunteers

#### Catheter placement/calibration

Prior to insertion, the solid-state sensor was calibrated according to the manufacturer’s instructions (see Additional file 1). Both catheters were inserted aimed at measuring Pes in the mid-esophageal range; location of the sensor corresponded to approximately halfway the balloon. Esophageal placement was confirmed by cardiac artifacts/esophageal spasms on the pressure waveforms. Balloon filling volume was checked with the Baydur maneuver (end-expiratory occlusion test) and adjusted when needed (see Additional file 1). The ΔPes_bal_/ΔPaw ratio was targeted at 0.9-1.1 for higher accuracy. If this range was not reached after five maneuvers, a range of 0.8-1.2 was accepted.

Measurements.

Recordings were obtained with subjects in sitting, semi-recumbent, supine and prone position. Balloon catheter accuracy was verified in between position changes. In each body position, 2 minutes of Pes during unloaded tidal breathing was acquired. During sitting and semi-recumbent position, subjects were additionally shortly exposed to three levels of inspiratory effort to obtain a variable within-subject range of effort (thus Pes values), using a threshold loading device (Power Breathe, POWERbreathe Ltd, UK), see Additional file 1.

### Patients

The study protocol was initiated directly after surgery upon arrival on the ICU. Ventilator settings were according to clinical protocols.

#### Catheter placement/calibration

Catheter positioning and calibration were similar as to healthy volunteers, while patients were still deeply sedated. Position was verified with thoracic X-ray when made within standard of care and/or using video laryngoscopy. The NutriVent catheter was initially used as comparator; however, in the first four patients, interference between both catheters was observed (see Results). From the 5^th^ patient, the Cooper catheter was used instead.

#### Measurements

During controlled ventilation, 10 minutes of tidal breathing were recorded, and three end-inspiratory and end-expiratory holds were performed (at 0, 5 and 10 minutes) for static measurements. Another 10-minute recording was performed during partially assisted ventilation when spontaneous breathing resumed as per clinical care. Correct balloon filling volume was verified at the start of each recording. Catheters were removed upon study completion.

### Offline analysis

For the bench study, the minimum, maximum and delta pressures were calculated using custom software (Polybench, Applied BioSignals, Germany). At each measurement time point, a median for each parameter was calculated over 60 preceding artificial breaths for further analyses.

For the healthy volunteers and patient studies, signal processing and analyses were performed in MATLAB (Mathworks, USA). Periods or individual breaths with substantial artifacts (e.g., esophageal spasms, coughing) were removed. Signals were processed using a 2^nd^-order 5 Hz low-pass Butterworth filter followed by a 0.1 second unweighted moving average filter. Static measurements for end-expiratory and end-inspiratory holds (in patients) and the Baydur maneuver (ΔPes_bal_/ΔPaw and ΔPes_solid_/ΔPaw) were manually selected from the tracings. Whereas the ΔPes_bal_/ΔPaw was used to verify balloon filling volume throughout the study, for the solid-state pressure transducer the Baydur test served as offline measure for sensor stability (as this catheter only requires zeroing before insertion). A breath detection algorithm was used and absolute values for end-inspiratory Pes, end-expiratory Pes and the resulting inspiratory amplitude (ΔPes) were computed breath-by-breath for both signals (in cmH_2_O).

### Endpoints

For the bench study, P_solid_ was compared to P_ref_. For healthy subjects and patients, the primary endpoint was the difference in absolute Pes values between the solid-state sensor and balloon catheter (Pes_solid_–Pes_bal_), measured at end-expiration and peak inspiration, and the difference in relative Pes values (i.e., inspiratory amplitude) between both catheters (ΔPes_solid_–ΔPes_bal_). Endpoints were separated for the different ventilation modes/populations. Secondary endpoints were the stability of the solid-state catheter as from repeated Baydur values, and device-related adverse events.

### Sample size

Sixteen catheters were tested on the bench; a convenience sample based on standard deviations (SD) obtained in the manufacturer’s previous technical tests. These tests were also used to substantiate sample sizes for the in-human studies, resulting in 7 subjects required for Bland-Altman analyses for each study, assuming a type-I error of 0.05 and type-II error of 0.20, and the following variables: expected mean (SD) of difference: 0.79 (0.53) cmH_2_O; maximum allowed difference between methods: 3.5 cmH_2_O, based on pressure accuracy tests and sensor drift tests (Pulmotech B.V.). We enrolled a larger sample (15 healthy subjects, 16 patients) to allow for more variability, to increase user-experience and to account for potential clinical/technical challenges.

### Statistical analysis

Statistical analyses were performed in R (RStudio, version 2024.02.2, Posit Software, PBC). For the bench, mean difference (i.e., P_solid_–P_ref_), and upper and lower limits of agreement (LoA) were calculated for each measurement time point. For the healthy volunteer and patient studies, baseline demographics and/or ventilator settings are presented as median (interquartile range, IQR) or numbers (%). Bland-Altman analysis with bootstrapping to account for variable number of breaths across subjects was performed to compare Pes values between catheters (see Additional file 1). For the healthy volunteers where variability in inspiratory effort was introduced with loaded breathing, agreement between Pes values for the solid-state vs. balloon was also evaluated with simple linear regression.

## RESULTS

### Bench study

The 16 solid-state sensors demonstrated a small positive bias (P_solid_–P_ref_) that increased from 0 to approximately 1.5 cmH_2_O during the first 10 hours for the absolute minimal and maximum pressures, and then remained stable at around 1 cmH_2_O until 120 hours (Figure 1AB). There was a negligible bias for pressure swings (0.13 cmH_2_O) throughout the full study (Figure 1C).

**Figure 1.**
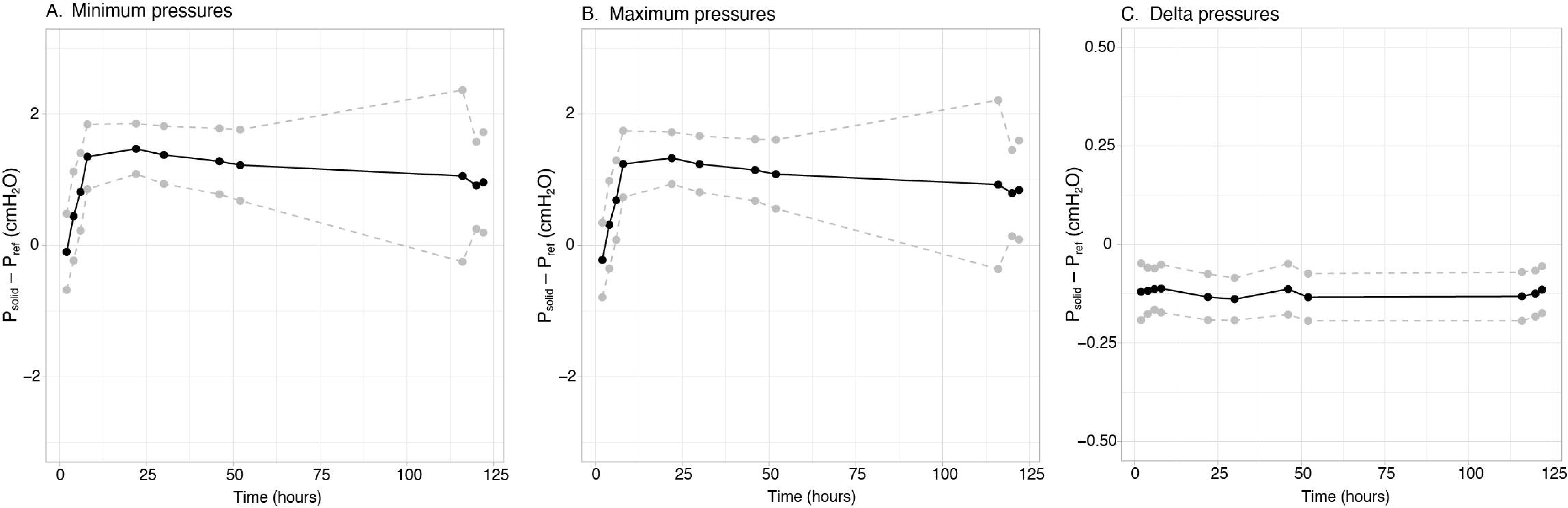
Bench results over a 120-hour measurement period, for the minimum pressures (A), maximum pressures (B) and delta pressures (C). Black line represents the mean difference (bias, i.e. solid-state catheter pressure (Psolid) minus reference pressure (Pref)), dashed gray lines are the upper and lower limits of agreement.

### Healthy volunteers

Fifteen healthy subjects (male/female 3/12; age 35.5±13.5 years) completed the study without adverse events. Two subjects were excluded from the full analysis, because of balloon catheter dislocation early in the study and inability to obtain reliable recordings after this was noticed (n=1), or balloon Baydur (ΔPes_bal_/ΔPaw) exceeding the 0.8-1.2 range throughout the study (n=1). For other subjects (n=5), short sections with low signal quality were removed (i.e., many esophageal spasms and/or cardiac artefacts, or ΔPes_bal_/ΔPaw not within 0.8-1.2 range for a specific body position).

### Comparisons

Figure 2 shows examples of Pes_bal_ and Pes_solid_ tracings. A total of 563 breaths of thirteen subjects were included in analyses when accepting tracings with ΔPes_bal_/ΔPaw within the 0.9-1.1 range. Bland-Altman analyses (Figure 3) revealed a bias (i.e., Pes_solid_ – Pes_bal_) [upper LoA; lower LoA] of 1.58 [8.19; −5.03], −2.37 [3.96; −8.69] and 3.94 [11.09; −3.20] cmH_2_O for end-expiratory, end-inspiratory and ΔPes values, respectively. Pes_bal_ and Pes_solid_ values showed moderate to good correlations (Additional files 4-6), with an average R^2^ of 0.70, 0.82 and 0.84 for end-expiratory, end-inspiratory and ΔPes values, respectively.

**Figure 2.**
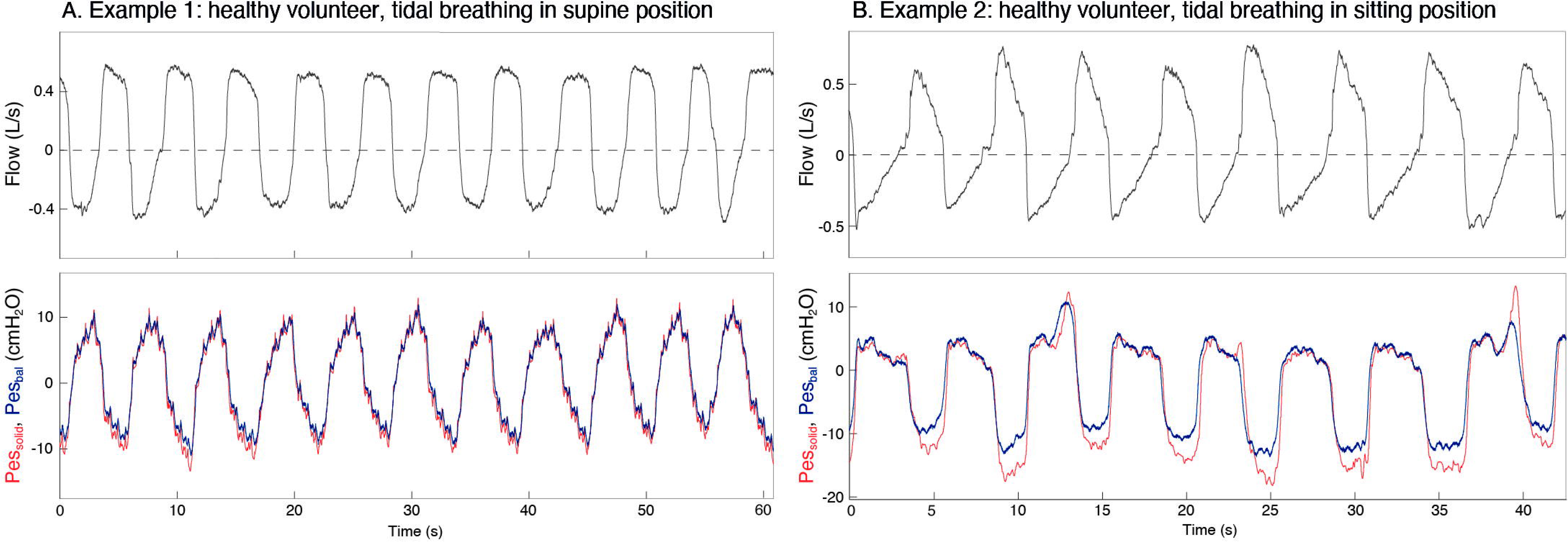
Examples of tracings obtained in two different healthy volunteers, during unloaded tidal breathing in supine (A) and sitting (B) position.

**Figure 3.**
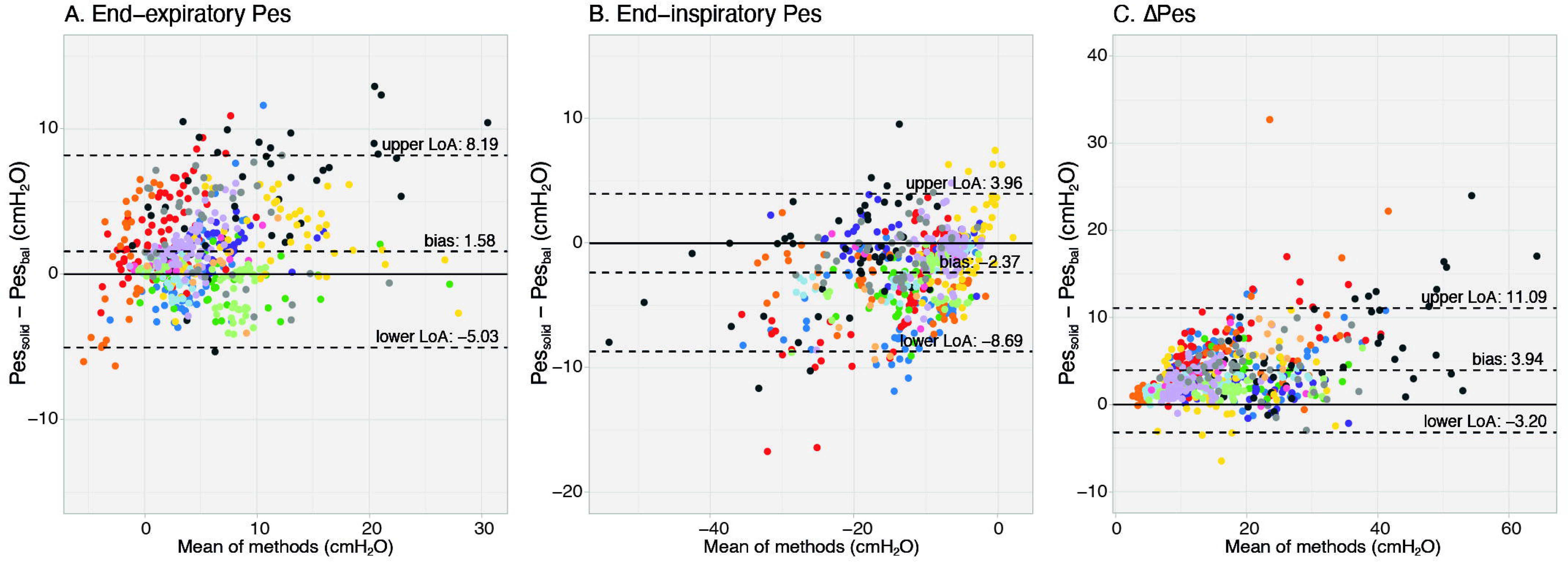
Bland-Altman results for healthy volunteers. Each color represents a different subject.

The average ΔPes_bal_/ΔPaw was 0.87±0.26 (min-max: 0.39-1.12; n=97 maneuvers), indicating that the balloon required frequent recalibrations after e.g. body position changes before adequate comparisons with Pes_solid_ could be made. For the solid-state catheter, Baydur maneuvers were analyzed offline to determine sensor stability. Of the 98 maneuvers analysed, ΔPes_solid_/ΔPaw ratio was 1.01±0.14 [min-max: 0.74-1.54]; 10 measurements (of which 4 from one subject) exceeded the 0.8-1.2 range.

We performed two sensitivity analysis: 1) when accepting tracings with ΔPes_bal_/ΔPaw within the 0.8-1.2 range, 877 breaths were included, resulting in comparable bias and LoA (Additional File 7); 2) when selecting only tracings with both ΔPes_solid_/ΔPaw and ΔPes_bal_/ΔPaw ratios between 0.9-1.1 and excluding one subject where cardiac artifacts amplitudes exceeded 5 cmH_2_O in both signals, 357 breaths from 11 subjects were used, resulting in a lower bias [upper LoA; lower LoA]: 0.68 [7.80; −6.44], −2.08 [3.21; −7.37] and 2.77 [9.42; −3.89] cmH_2_O for end-expiratory, end-inspiratory and ΔPes values, respectively.

### Patients

27 patients consented prior to their surgery, and 16 patients (see Table 1 for characteristics) were eventually enrolled upon ICU arrival. Reasons for withdrawal were a last-minute canceled/rescheduled surgery (n=10) and hemodynamic instability (n=1). One patient was excluded from the full analysis, as the solid-state sensor demonstrated non-physiological signals (i.e., Pes swings exceeding Paw) and the Nutrivent catheter was unreliable due to balloon emptying despite recalibration attempts (Additional file 8). For controlled ventilation, one additional patient was excluded due to balloon (Cooper) catheter emptying (but included for assisted ventilation after adequate recalibration). For assisted ventilation, three additional patients were excluded (but included in controlled ventilation analysis) due to: technical issues (n=1), very low breathing efforts (n=1), many artifacts hampering breath detection (n=1). This resulted in a total analysis set of 2200 breaths from 14 patients during controlled ventilation and 889 breaths from 12 patients during assisted ventilation; all Pes_bal_ tracings were adequately calibrated with a Baydur maneuver between 0.9-1.1. No adverse events were reported.

**Table 1.** Main characteristics of the study population.

| Characteristic | Total (N = 16) |  |
| --- | --- | --- |
| Age, years; median (IQR) | 65 (58 - 69) |  |
| Male, sex; n (%) | 15 (94) |  |
| BMI kg/m<sup>2</sup>; median (IQR) | 26.7 (25.0 - 29.7) |  |
| IBW, kg; median (IQR) | 76.9 (68.1 - 79.2) |  |
| Medical history (n) |  |  |
| Asthma | 1 |  |
| Non-obstructive emphysema | 1 |  |
| Coronary artery disease | 6 |  |
| Renal Failure | 2 |  |
| Type of surgery performed (n) |  |  |
| CABG | 5 |  |
| CABG + AVR | 2 |  |
| ROSS procedure | 2 |  |
| MVP-MIC | 3 |  |
| AVR | 4 |  |
|  | Controlled ventilation<br>(median (IQR)) | Assisted ventilation<br>(median (IQR)) |
| Respiratory parameters |  |  |
| PEEP set (cmH<sub>2</sub>O) | 7 (7 - 8) | 7 (6 - 8) |
| PEEP total (cmH<sub>2</sub>O) | 8.5 (7.7 - 9.1) | N/A |
| PC/PS above PEEP (cmH<sub>2</sub>O) | 11 (10 - 13.3) | 7 (5 - 8) |
| Pplat (cmH<sub>2</sub>O) | 17 (16.2 - 19.7) | N/A |
| Driving Pressure (cmH<sub>2</sub>O) | 9.5 (7.7-10.7) | N/A |
| Pocc (cmH<sub>2</sub>O) | N/A | 12.7 (8.4 - 17.3) |
| RR (/min) | 16 (16 - 18) | 13 (9 - 15) |
| Tidal Volume (mL) | 484 (448 - 568) | 689 (459 - 856) |
| Minute Volume (L/min) | 7.9 (7.0 - 8.7) | 6 (5.6 - 7.3) |
| FiO<sub>2</sub> | 30 (30 - 41) | 35 (30 - 41) |
| SpO<sub>2</sub> | 98 (97 - 98) | 98 (97 - 99) |
| EtCO<sub>2</sub> | 5.4 (4.6 - 5.9) | 6.8 (5.8 - 7.3) |
| Hemodynamic parameters |  |  |
| Heart rate (/min) | 75 (68 - 81) | 82 (79 - 90) |
| BP systolic (mmHg) | 103 (94 - 109) | 138 (122 - 154) |
| BP diastolic (mmHg) | 56 (53 - 59) | 65 (61 - 68) |
| MAP (mmHg) | 70 (66 - 74) | 85 (80 - 95) |

*Abbreviations: AVR aortic valve replacement, BMI body mass index, BP Blood pressure, IBW ideal body weight, CABG coronary artery bypass graft, EtCO<sub>2</sub> End-tidal CO<sub>2</sub>, FiO<sub>2</sub> fraction of inspired oxygen, IQR interquartile Range, MAP Mean Arterial Pressure, MVP mitral valve repair, MIC minimal invasive surgery, PEEP positive end-expiratory pressure, PC pressure control, PS Pressure Support, Pplat plateau pressure, Pocc occlusion pressure, RR Respiratory Rate, ROSS procedure aortic valve replacement with own pulmonary valve and pulmonary allograft, SpO<sub>2</sub> oxygen saturation*

### Signal interference

Signal interference between the solid-state and Nutrivent catheter is explained in Additional file 9, likely the result of having two rather thick catheters (with large balloon of Nutrivent catheter) in place and rendering parts of the data unusable for further analysis. After using the Cooper catheter (from the 5^th^ patient), such interference was not observed.

### Comparisons

Figure 4 shows Pes_bal_ and Pes_solid_ tracings during controlled and assisted ventilation. During controlled ventilation, Bland-Altman analyses (Figure 5A-C) revealed a low bias (i.e., Pes_solid_ – Pes_bal_)) [upper LoA; lower LoA] of −0.15 [1.39; −1.70], 0.32 [3.35; −2.72] and 0.47 [3.79; −2.85] cmH_2_O for end-expiratory, end-inspiratory and ΔPes values, respectively. Patient 16 demonstrated inspiratory pressure amplifications in Pes_solid_ that we could not attribute to cardiac artifacts solely (Additional file 10). Removing this patient improved comparisons: bias [upper LoA; lower LoA] of −0.08 [1.43; −1.58], 0.07 [2.62; −2.49] and 0.15 [2.63; −2.34] cmH_2_O for end-expiratory, end-inspiratory and ΔPes values, respectively.

**Figure 4.**
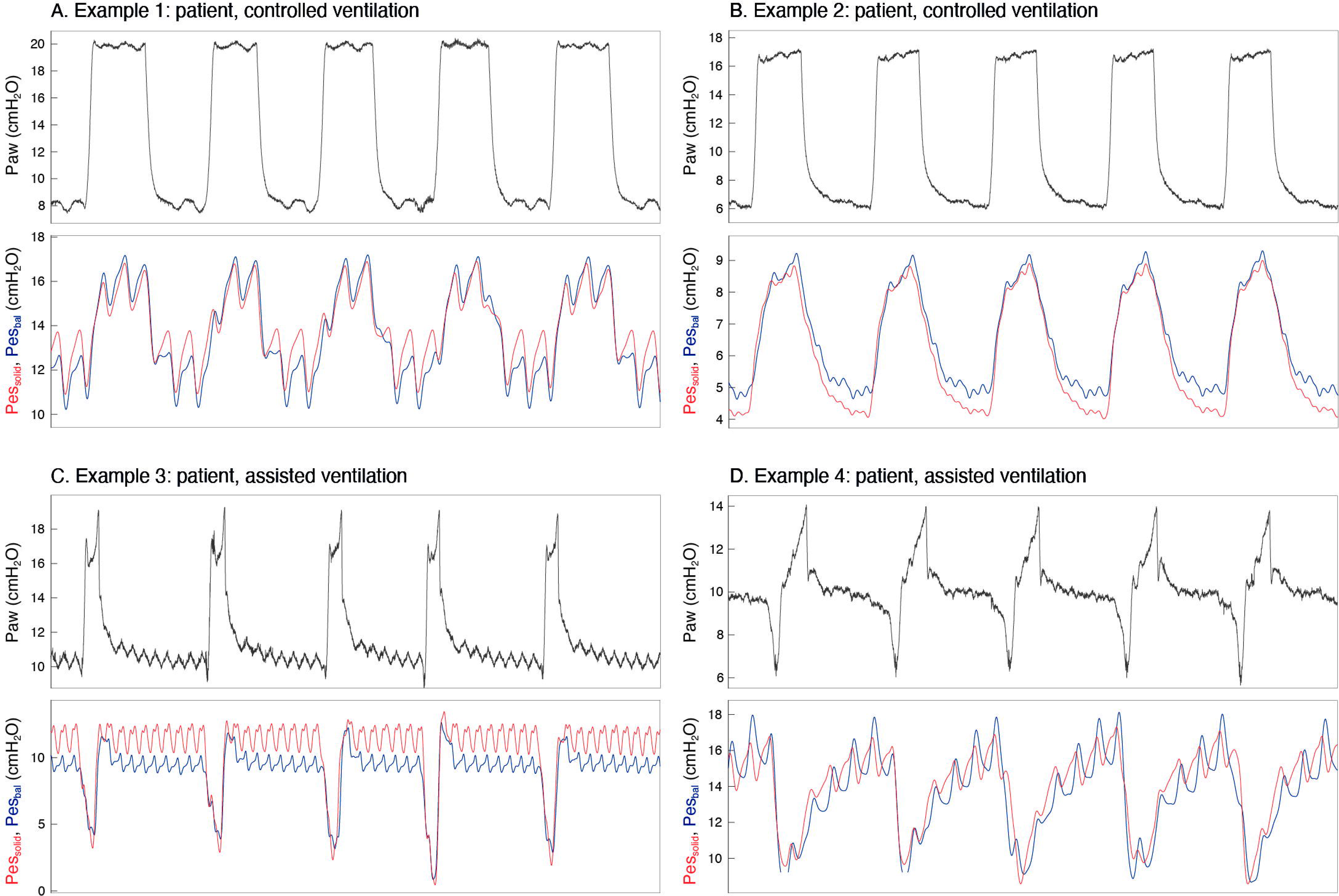
Examples of tracings obtained in four different patients during controlled ventilation (A, B) and assisted ventilation (C, D).

**Figure 5.**
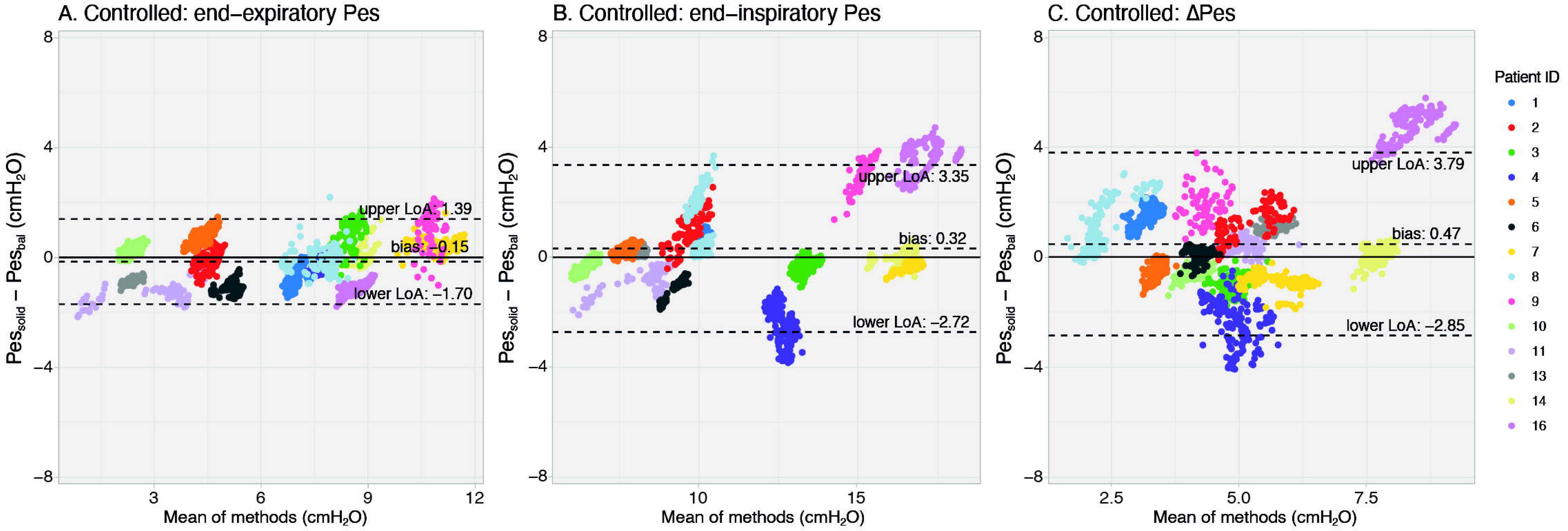
Bland-Altman results for patients on controlled ventilation. Each color represents a different patient.

Pressures obtained during static conditions are presented in Table 2; LoA were smaller (all <2 cmH_2_O) as compared to breath-by-breath analysis.

**Table 2.** Static comparisons for Pes_solid_ – Pes_bal_ (controlled ventilation)^1.^

|  | Expiratory hold | Inspiratory hold | ΔPes |
| --- | --- | --- | --- |
| Mean difference (cmH<sub>2</sub>O) | -0.21 | -0.36 | -0.15 |
| SD of difference (cmH<sub>2</sub>O) | 0.90 | 0.77 | 0.76 |
| Upper LoA (cmH<sub>2</sub>O) | 1.56 | 1.16 | 1.33 |
| Lower LoA (cmH<sub>2</sub>O) | -1.98 | -1.87 | -1.63 |

<sup>1</sup>Patient 16 was removed from this analysis due to unphysiological inspiratory and delta Pes values.

During assisted ventilation, bias remained low, but LoAs were wider, yet smaller than in healthy volunteers (Figure 6): −0.20 [5.02; −5.41], −0.54 [4.60; −5.68] and 0.35 [3.88; −3.18] cmH_2_O for end-expiratory, end-inspiratory and ΔPes values, respectively.

**Figure 6.**
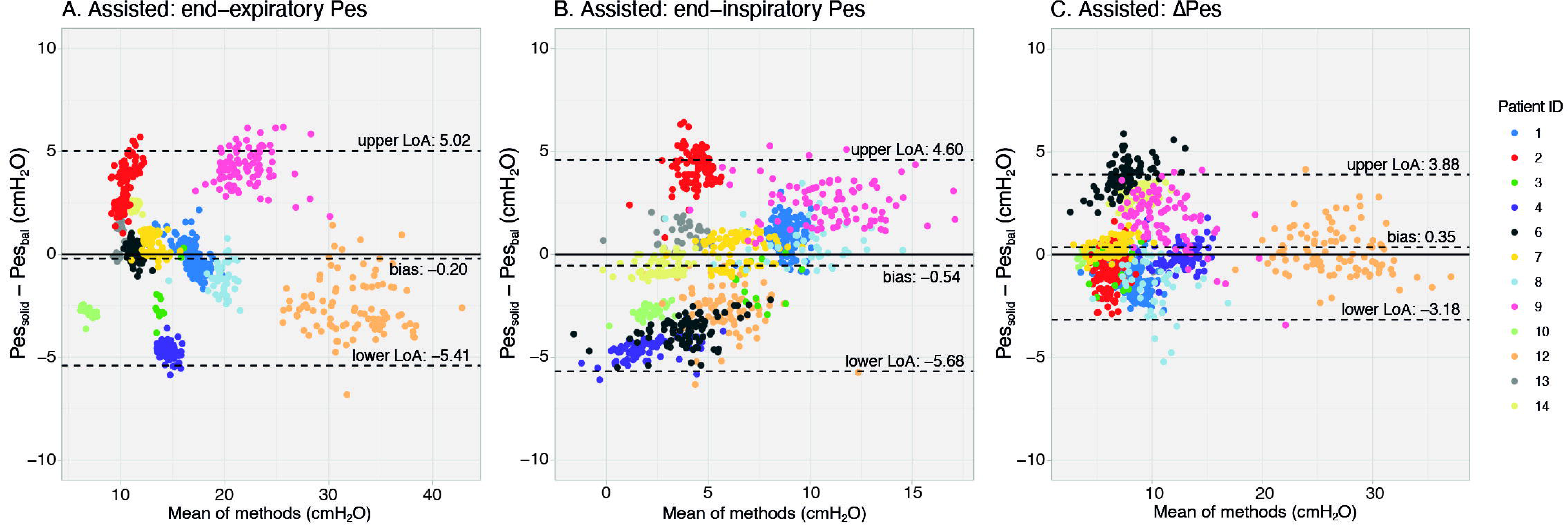
Bland-Altman results for patients on assisted ventilation. Each color represents a different patient; subject’s color coding is similar for Figure 5.

A total of 24 Baydur maneuvers from the solid-state catheter were analyzed to determine sensor stability (for n=12 patients in controlled ventilation (2 missing as acquisition started after balloon calibration) and for n=12 patients in assisted mode). The mean ΔPes_solid_/ΔPaw ratio was 1.05±0.18 [min-max: 0.72-1.48].

## DISCUSSION

We tested a new Pes catheter with solid-state sensor on the bench, in healthy volunteers and in ventilated patients. Findings can be summarized as: 1) the solid-state sensor demonstrated excellent agreement with a reference pressure during a 5-day bench test, without signal drift; 2) moderate to good agreements with Pes_bal_ during tidal breathing was found in healthy volunteers when Pes_bal_ was adequately calibrated; 3) these agreements improved in ventilated patients during tidal breathing (controlled and assisted ventilation) and further in static conditions (breath holds); 4) the sensor remained stable throughout study recordings; 5) the solid-state Pes catheter often measured higher ΔPes values, especially in healthy volunteers during high efforts.

### Validity of reference pressure

For our in-human comparisons, balloon Pes catheters served as “reference standard” as the true pleural pressure is not available in humans. A concern with this approach in our and previous studies[6–9] is that each balloon has an optimal filling volume, depending on its perimeter/length, and elasticity and length of connecting tubing. These characteristics, but also the balloon’s position in the esophagus and ex-vivo pressure sensor affect absolute pressure values and the balloon’s capacity to respond to Pes swings (i.e., frequency response)[10]. Furthermore, the balloon may empty over time and recommended filling volumes by manufacturers are often not optimal clinically; changes in intrathoracic pressure or chest wall compliance (e.g. change in PEEP, body position, pleural pressure inhomogeneities) require recalibration[1,3,10,11]. Indeed, uncalibrated and calibrated balloon pressures (i.e., corrected for esophageal wall and balloon elasticity) could differ at end-expiration by 5.1 cmH_2_O (range: 0.8 to 35.1 cmH_2_O) despite obtaining a Baydur range of 0.8-1.2.[10] This makes the position of balloon catheters as reference standard somewhat questionable. We aimed to target higher balloon catheter accuracy for primary comparisons (ΔPes_bal_/ΔPaw within 0.9-1.1 range) to improve comparisons, but this was sometimes challenging in healthy volunteers. A Baydur range of 0.8-1.2 is considered acceptable in literature[1] but also implies an accepted deviation of 20% from the true ΔPes value, which may impact clinical decision-making when applying Pes-based ventilation strategies. Considering the solid-state sensor stability, represented by the excellent offline obtained ΔPes_solid_/ΔPaw (1.01±0.14 in healthy volunteers, 1.05±0.18 in patients) and bench results, it can also be argued that the solid-state sensor better represented the actual (delta) Pes. Studies comparing different Pes sensor types (i.e., solid-state, balloons [11], liquid-filled catheters) are therefore challenging to interpret without the clinical availability of a true reference standard.

### Related works

Over 20 years ago, solid-state Pes catheters were compared with a balloon catheter in healthy volunteers[6,8] demonstrating reliable relative/delta Pes values. However, uncontrollable offset shifts (10 cmH_2_O for transpulmonary pressure[6]) and a high bias (>7 cmH_2_O[8]) were observed, making absolute values unreliable. Authors hypothesized that Van der Waal forces contributed to falsely high pressures (e.g., mucus sticking to sensor membrane, or contact with the esophageal wall)[6] and negative signal drifts[8]. More recent work in 2017[7] and 2021[9] comparing micro-transducers with balloon catheters report a smaller bias: end-expiratory Pes of −3.6 cmH_2_O[7] (vs. 1.6 cmH_2_O in our study) and delta Pes of 3.8 cmH_2_O[9] (vs. 3.9 cmH_2_O in our study), respectively.

The smaller[6, 8] or comparable[7, 9] biases in our healthy volunteers can be explained as follows. First, our solid-state catheter includes a small balloon above the sensor serving as a stabilizer to avoid sensor sticking to the esophageal wall; this likely also kept mucus off the sensor membrane, avoiding signal drifts. Second, the sensor is both temperature and humidity calibrated. Yet, in some subjects/patients large differences with Pes_bal_ were found, which may be explained by the sensor’s fast frequency response: since pressures are measured directly inside the esophagus signal dampening is avoided, but artifacts can be easily amplified. Cardiac artifacts in Pes_solid_ were sometimes high and body position dependent (e.g., more marked in supine position, see Additional file 11, in line with[8]), and more negative Pes values were observed with larger inspiratory efforts (Figure 2). This warrants careful identification and interpretation of artifacts, improved signal filtering at the bedside, and/or optimizing the sensor positioning. The smaller biases and LoA in ICU patients as compared to healthy volunteers could be explained by the stable ventilator settings and the low variability in breathing efforts during assisted mode. In addition, body position alterations were not part of the patient study.

### Strengths and limitations

This is the first study validating a novel solid-state Pes catheter, combining bench work with measurement in healthy volunteers and ICU patients during different ventilation modes. The use of the solid-state Pes catheter was considered easy: it requires only one calibration prior to insertion and pressures are measured directly in the esophagus – hence, some secondary limitations of balloon catheters (e.g., need for precise filling volume, risks of balloon emptying over time) are not applicable. In contrast, with the sensor’s fast response time, there is a possibility of pressure amplifications related to e.g., cardiac artifacts as discussed above. Our study also has some limitations. First, we used two different balloon catheters in the patient study, which was initially designed with the Nutrivent catheter as comparator, but significant interference was observed. Second, several short sections were excluded from the analysis due to artifacts in both signals and/or unreliable Pes_bal_ tracings. Nevertheless, when selecting stable tracings where the balloon was properly calibrated, good agreement between Pes_bal_ and Pes_solid_ was found. Since our sample size was larger than necessary, excluding some tracings likely did not affect power of our analyses. Third, in healthy volunteers we measured positive and high end-expiratory Pes values, likely the result of inspiratory effort maneuvers and/or cardiac artifacts. In addition, two patients demonstrated unphysiologically high Pes_solid_ values (see Results) despite proper calibration, reposition attempts and verification of esophageal positioning. Last, we did not perform multiple-day testing of the solid-state catheter in the ICU, but extensive bench tests demonstrated only minimal signal drift over 5 days.

### Clinical relevance

Pes monitoring allows individualization of ventilator settings via a more thorough understanding of the mechanical properties of the respiratory system. Over the last years, use of the balloon catheters has improved with the availability of dedicated monitors or ventilator connections. Yet, measurements remain technically challenging and widespread routine implementation is lacking. The solid-state catheter requires only one calibration prior to insertion, contributing to its ease of use. This makes the technique interesting for future implementation, also in the light of the recent regulatory challenges such as the medical device regulation in the European Union, which has put extensive pressure on the production of medical devices, resulting in limited availability and even withdrawal of certain balloon catheters from the market. Future work should focus on the longer-term use of the technique, e.g., in multiple-day measurements within a Pes-guided ventilation strategy, and optimizing filtering of (cardiac) artifacts and (automated) signal quality checks. In conclusion, this is the first study validating a novel solid-state Pes catheter in vitro, in healthy volunteers and in postoperative mechanically ventilated ICU-patients, with promising results. This could contribute to the implementation of Pes as advanced respiratory monitoring technique.

## DECLARATIONS

### Ethics approval and consent to participate

This study was approved by the local medical ethics committee of the Vrije Universiteit medical center (METc 2020.470) and Erasmus Medical Center (MEC-2023-0119). All healthy subjects and patients provided written informed consent prior to participation.

### Consent for publication

Not applicable

### Availability of data and materials

The datasets used and/or analyzed during the current study are available from the corresponding author upon reasonable request.

## Competing interests

RF and RS are employed by Pulmotech B.V. AJ and LH received research funding (paid to the institution) of Pulmotech B.V. for the conduct of the study.

## Funding

The study was partially funded by a grant from Pulmotech B.V., Leek, The Netherlands.

## Authors contributions

Concept: AHJ, RF, LH; Design: LH, AHJ; Data acquisition bench study: RF, RS; Data acquisition healthy volunteers: AM, LH, AHJ; Data acquisition patients: JPvO, NG, RF, RS, AHJ; Data analysis: JPvO, NG, AM, RF, MF, LH, AHJ; Data interpretation: all authors; Manuscript drafting: JPvO, NG, AHJ; Supervision: LH, AHJ; Manuscript revising for intellectual content and final approval: all authors.

## Data Availability

All data produced in the present study are available upon reasonable request to the authors

## Acknowledgements

We thank all healthy volunteers and patients that were willing to contribute to the study.

## LIST OF ABBREVIATIONS

BMI: Body Mass Index
ICU: Intensive Care Unit
iEPC: Intelligent Esophageal Pressure Catheter
iEPMS: Intelligent Esophageal Pressure Monitoring System
LoA: Limit of Agreement
Paw: Airway pressure
Pes: Esophageal pressure
Pes_bal_: Esophageal pressure measured by balloon catheter
P_ref_: Reference pressure
P_solid_: Esophageal pressure measured by the solid-state pressure sensor

## Notes

### Clinical Trial

NCT05817968

