## Additional file 1 for "Solid-state esophageal pressure sensor for the estimation of pleural pressure: a bench and first-in-human validation study"

*Additional file 1 to:*

Annemijn H. Jonkman^1^

**Author affiliations:**

1) Intensive Care, Erasmus Medical Center, Rotterdam, The Netherlands
2) Intensive Care, Amsterdam UMC location Vrije Universiteit Amsterdam, The Netherlands
3) Amsterdam Cardiovascular Sciences, Amsterdam, the Netherlands
4) Pulmotech B.V., Leek, The Netherlands
5) Intensive Care, Radboud University Medical Center, Nijmegen, The Netherlands

Calibration of the solid-state Pes catheter

Prior to insertion and when connected to the acquisition device, the solid-state sensor was calibrated: two layers of sterile gauze pad soaked in water were placed on the sensor to create a humid environment. After 2 minutes the zero button was pressed and it was verified that pressures were 0 cmH_2_O.

***Healthy volunteers***

Baydur test

Subjects were breathing through a flow-sensor connected to a mouthpiece while wearing a nose clip. One limb of the flow sensor (differential pressure) was used to acquire airway pressure. The distal part of the flow sensor should be occluded during the Baydur test. This can be done e.g., with a hand, or using a loading device with maximum resistance (fully occluded) as illustrated below. An example of a Baydur measurement is shown (ΔPes_bal_/ΔPaw = 1.08).

| 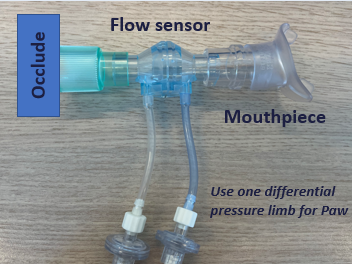 | 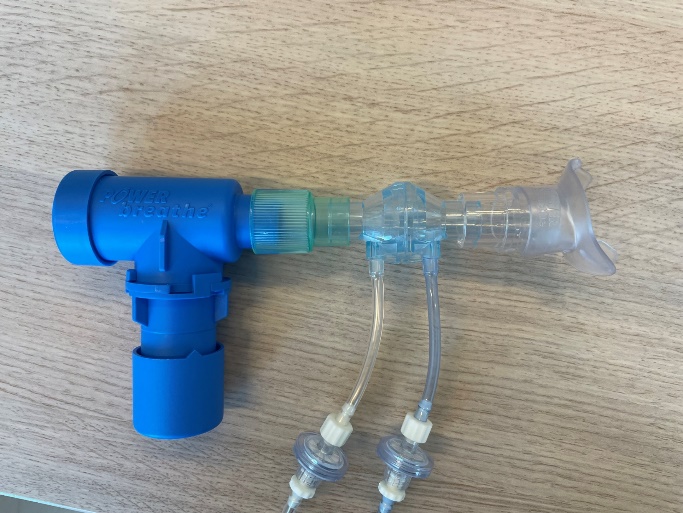  Occluded threshold loading device |
| --- | --- |
| 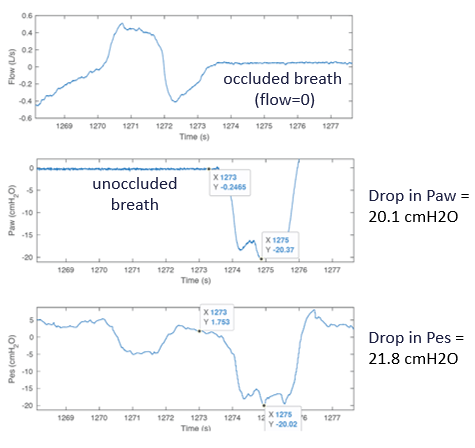 | |

**Additional figure 1.** The loading device to perform a Baydur test in healthy volunteers with an example of a Baydur measurement.

***Inspiratory loading protocol***

During sitting and semi-recumbent position, subjects were additionally exposed to three levels of inspiratory effort to obtain a variable within-subject range of effort and thus Pes values: medium loading (30% of maximum inspiratory pressure (PImax)), high loading (60% PImax) and maximum loading (90% of PImax). PImax was assessed prior to study recordings with maximum inspiratory effort maneuvers. Loading was applied by letting the volunteer breathe through a mouthpiece connected to a threshold loading device (Power Breathe, POWERbreathe Ltd, UK) while wearing a nose-clip. The duration of the loading task was decreased as load increased: 2 minutes quiet breathing – 45 seconds medium loading – 1 minute rest – 30 seconds high loading – 1 minute rest – 15 seconds maximum loading.

***Statistics: Bland-Altman analysis***

Bland-Altman analysis was performed comparing breath-by-breath Pes values between catheters. To correct for different numbers of measurements between subjects (e.g. due to varying respiratory rates), nonparametric bootstrap resampling was performed with 1000 iterations to compute the 95% confidence interval (CI) of lower and upper LoA. The same number of breaths, corresponding to the median number of breaths across all patients, was randomly selected for each patient during the resampling process. For the static measurements in patients (three end-expiratory and end-inspiratory holds during controlled ventilation), mean difference, SD of differences and 95% lower and upper LoA were computed.
