## Additional file 2 for "Solid-state esophageal pressure sensor for the estimation of pleural pressure: a bench and first-in-human validation study"

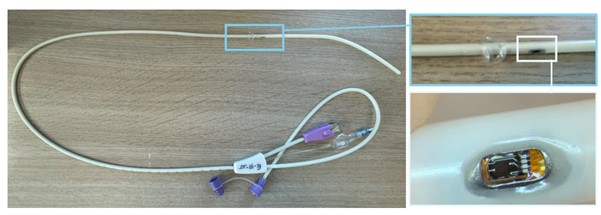

**Additional figure 2.** Intelligent esophageal pressure catheter (iEPC). The tiny solid-state sensor is embedded in the catheter, with a small balloon just above this sensor (top right photo). This balloon does not record any pressures but serves as a placeholder to make sure the sensor does not attach/stick to the esophageal wall.
