## Supplementary figures and images for "Solid-state esophageal pressure sensor for the estimation of pleural pressure: a bench and first-in-human validation study"

### Additional file 3

**Additional file 3**

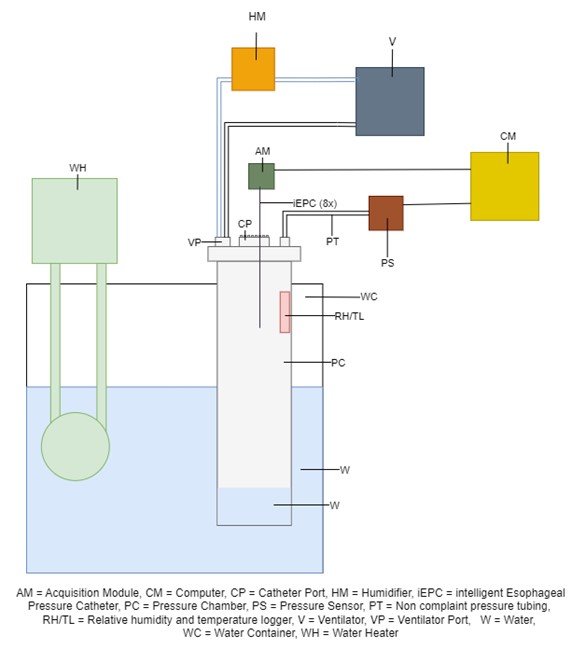

**Additional figure 3.** Bench setup for 5-day testing.
