## Additional file 5 for "Solid-state esophageal pressure sensor for the estimation of pleural pressure: a bench and first-in-human validation study"

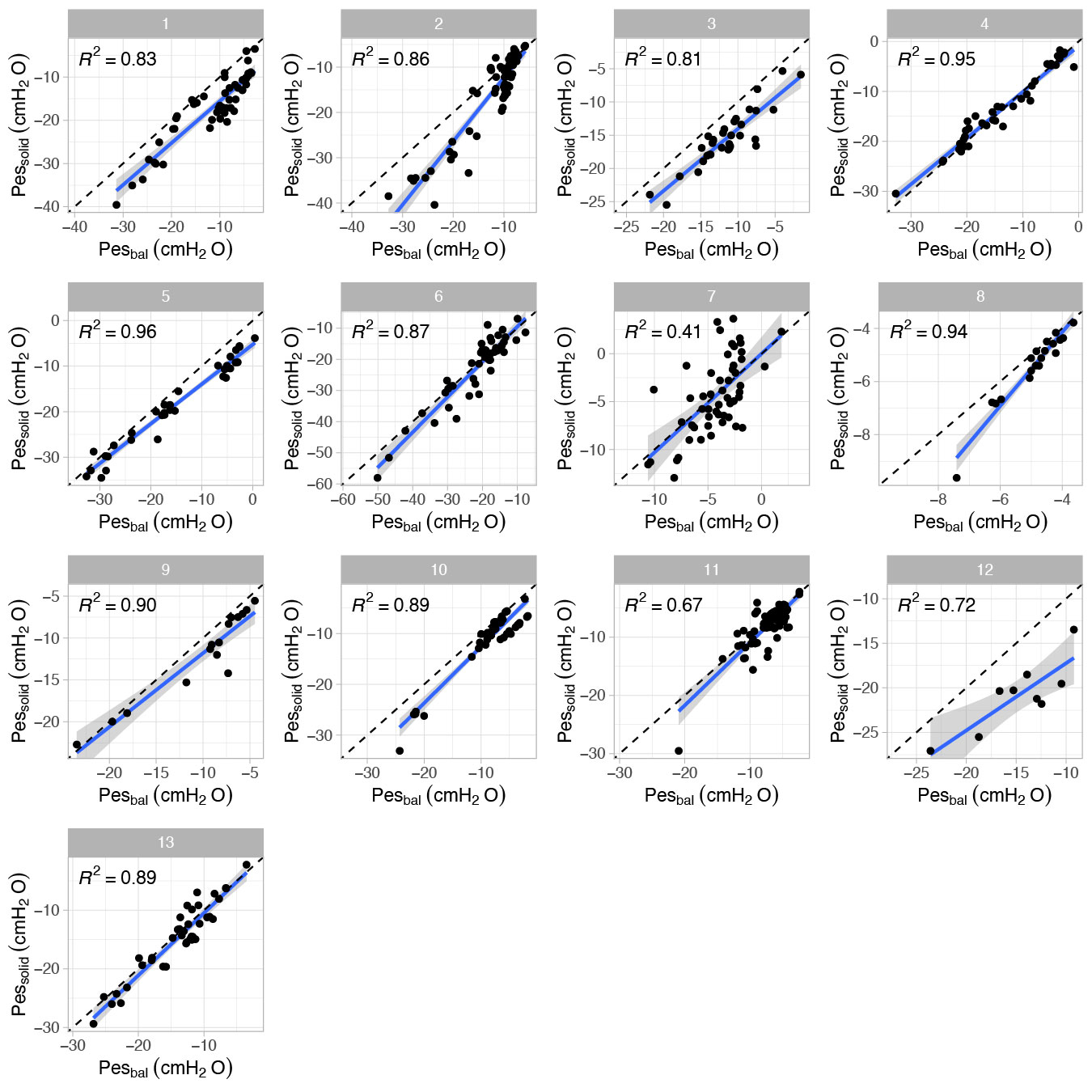

**Additional figure 5.** Healthy volunteers: regression analysis for Pes values at end-inspiration separated per subject.
