## Additional file 6 for "Solid-state esophageal pressure sensor for the estimation of pleural pressure: a bench and first-in-human validation study"

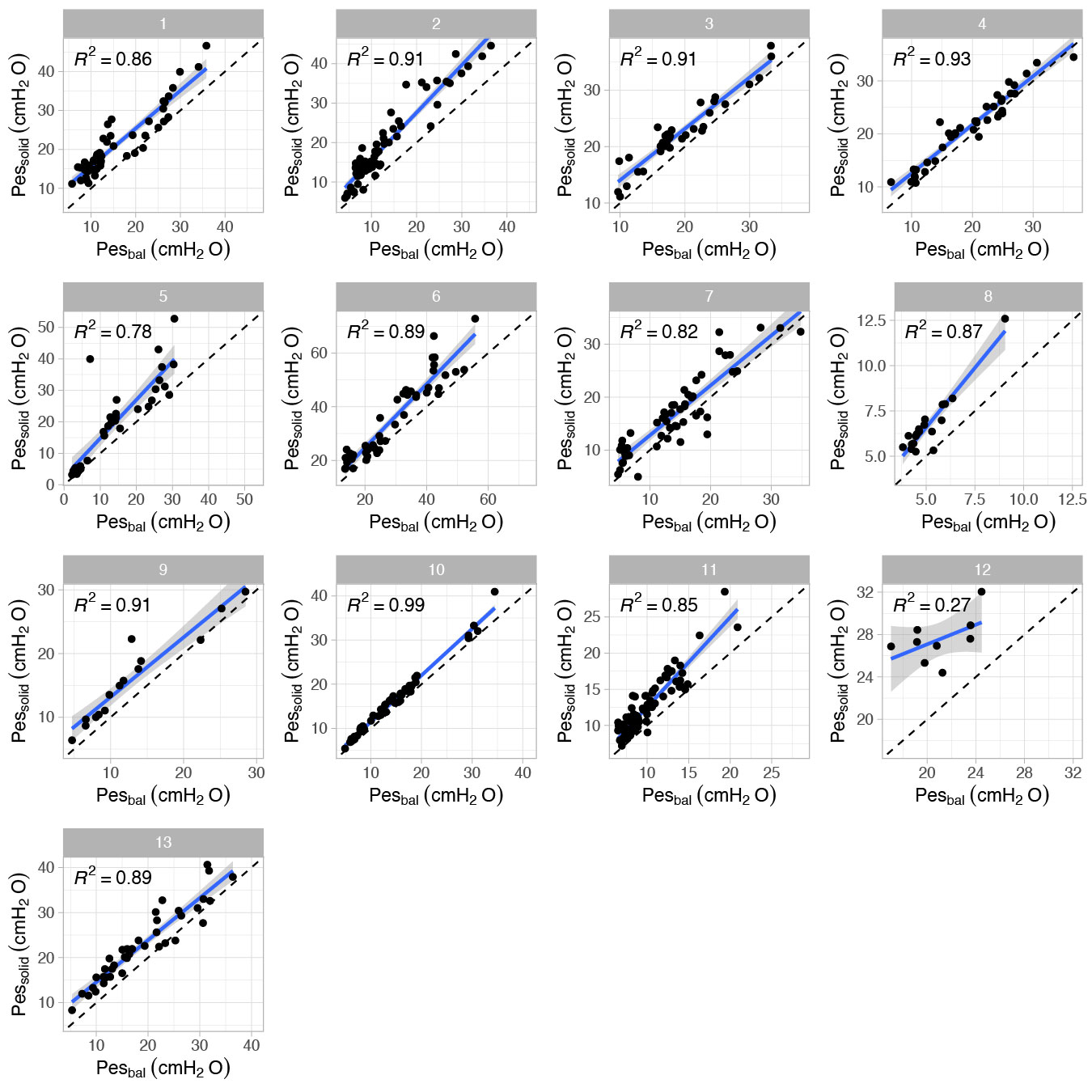

**Additional figure 6.** Healthy volunteers: regression analysis for ∆Pes values separated per subject.
