## Additional file 7 for "Solid-state esophageal pressure sensor for the estimation of pleural pressure: a bench and first-in-human validation study"

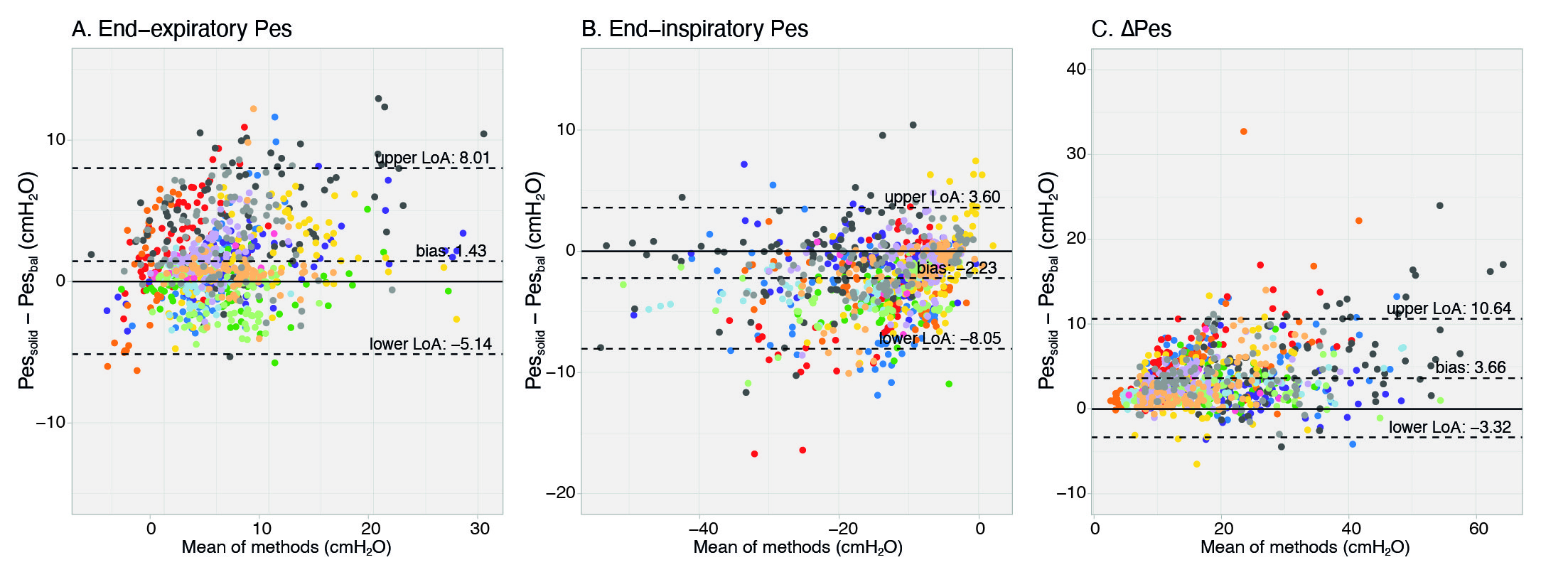

**Additional figure 7.** Healthy volunteers: Bland-Altman results for Baydur balloon within range 0.8-1.2. Each color represent a different subject.
