## Additional file 8 for "Solid-state esophageal pressure sensor for the estimation of pleural pressure: a bench and first-in-human validation study"

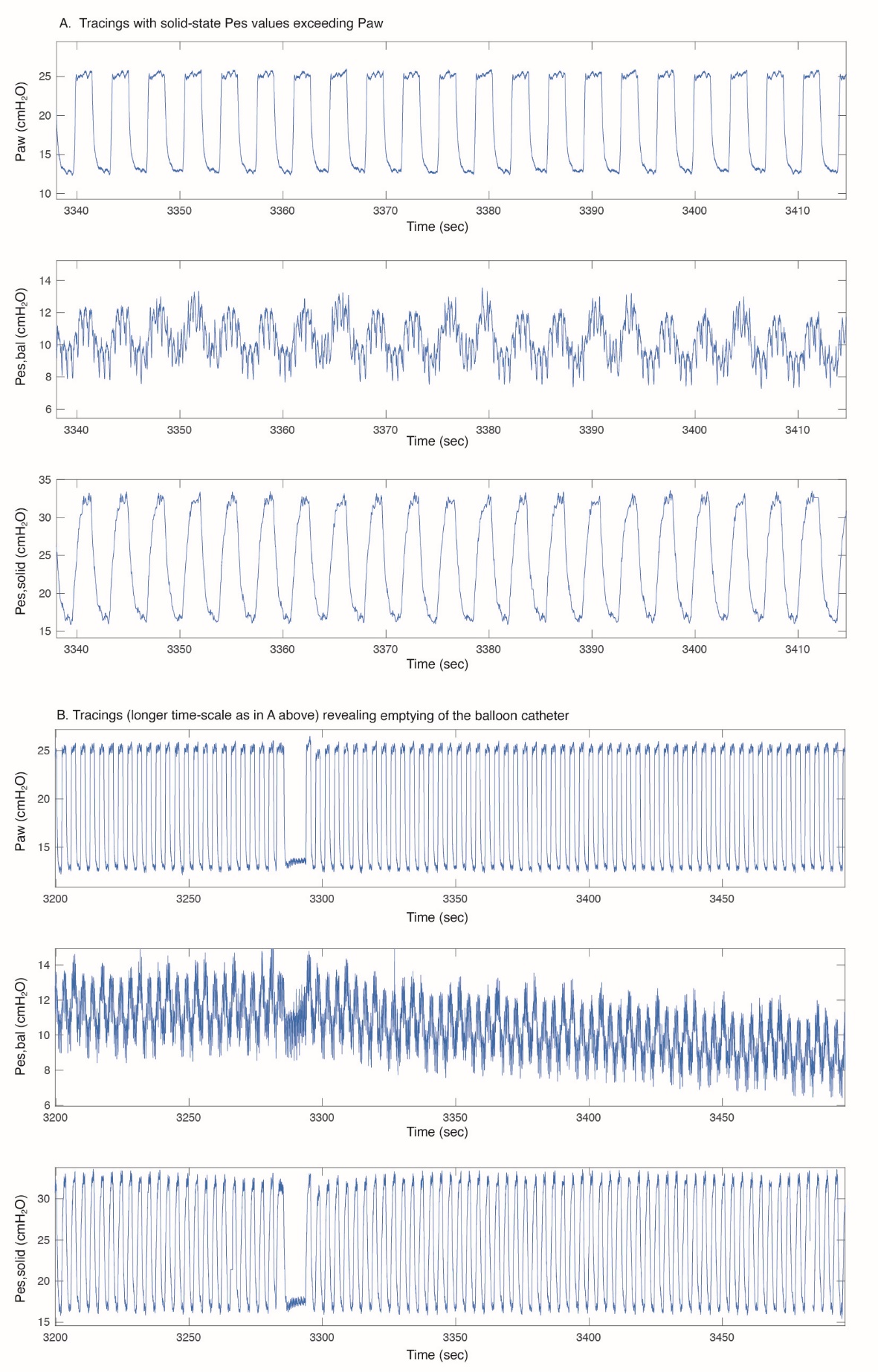

**Additional figure 8.** A. Signals from excluded patient as the solid-state sensor demonstrated non-physiological signals (i.e., Pes swings exceeding Paw). B. At the same time, the balloon catheter was unreliable due to emptying despite several recalibration attempts.
