## Additional file 9 for "Solid-state esophageal pressure sensor for the estimation of pleural pressure: a bench and first-in-human validation study"

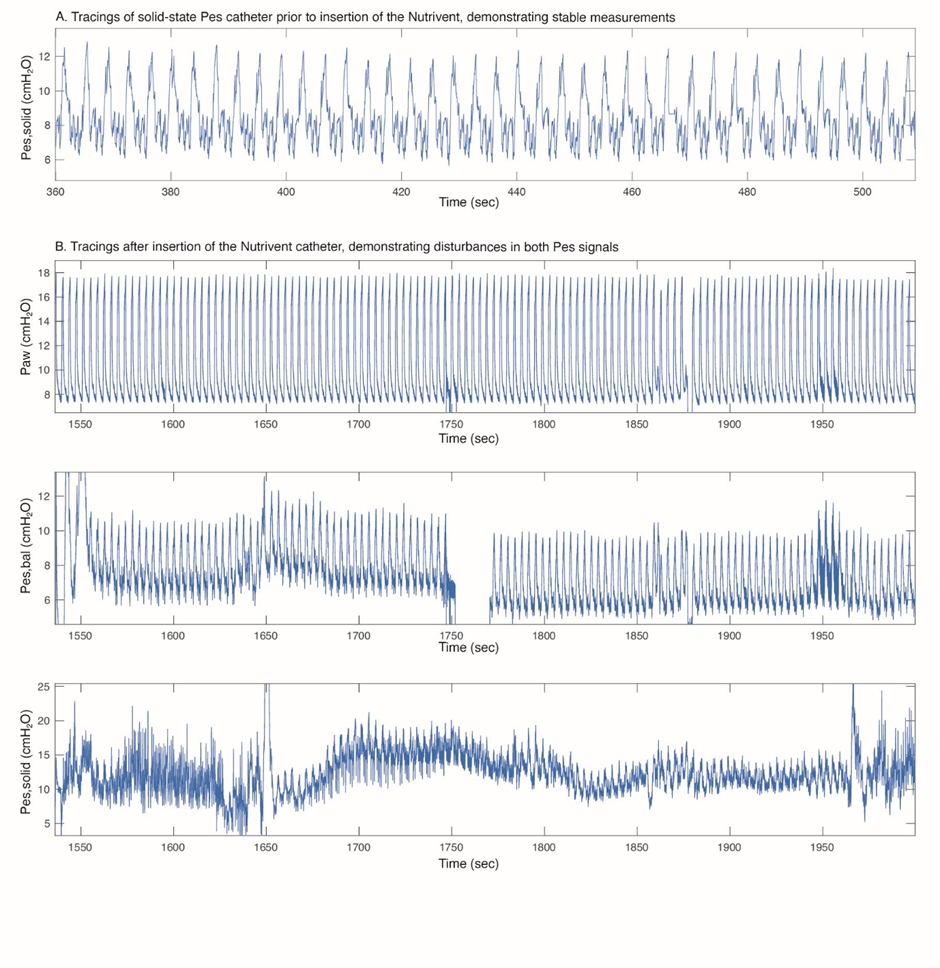

**Additional figure 9.** Signal interference between the solid-state and Nutrivent catheter. While Pes_solid_ demonstrated adequate signals prior to insertion of the balloon catheter (A), tracings became unstable afterwards (B). After several repositioning/refilling attempts of both catheters some stable parts were used for comparisons (not shown in this figure), despite artefacts in Pes_solid_. At 1750 seconds the balloon catheter briefly disconnected from the acquisition system (gap in recording).
