## Additional file 10 for "Solid-state esophageal pressure sensor for the estimation of pleural pressure: a bench and first-in-human validation study"

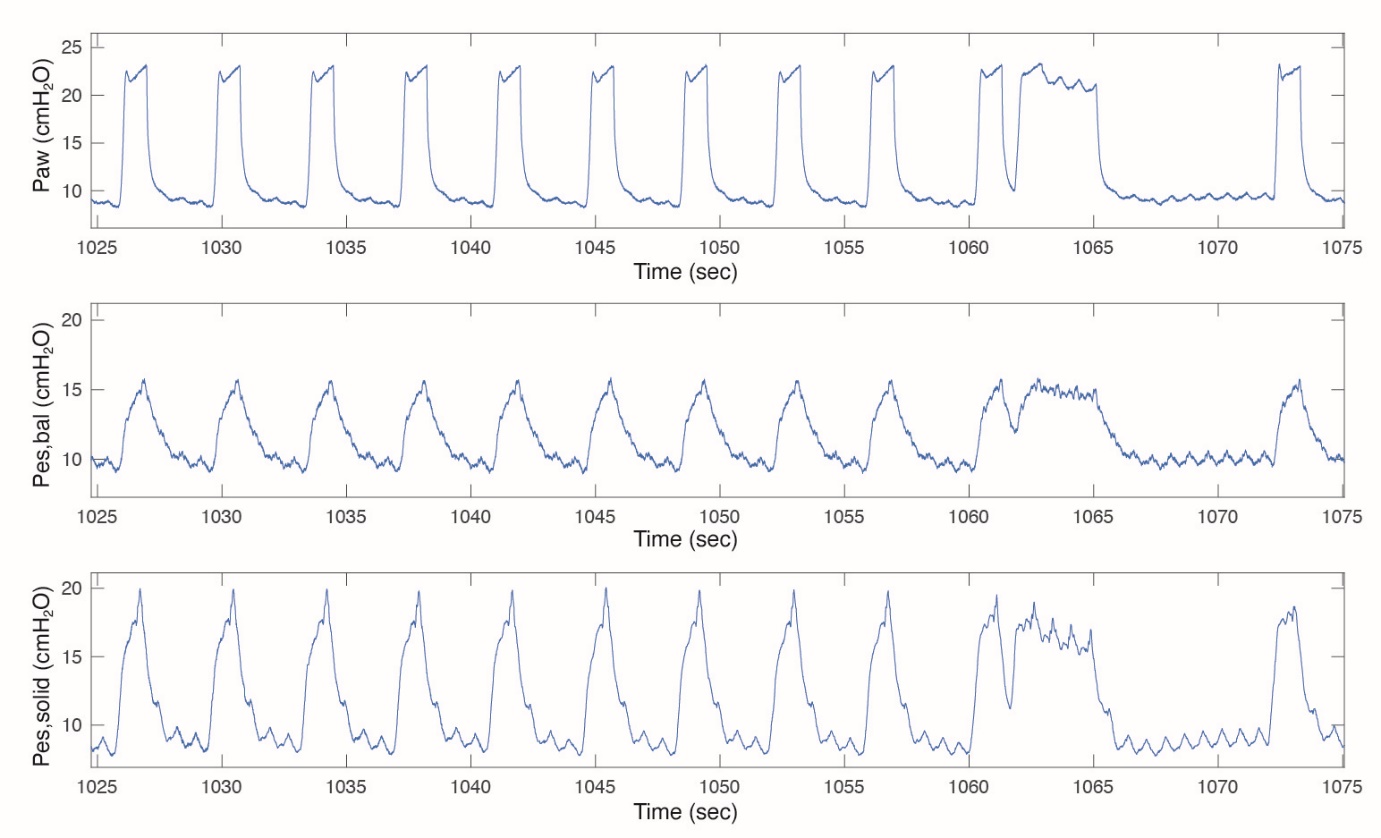

**Additional figure 10.** Example of a patient with very high inspiratory and thus also high ΔPes_solid_ values that we could not attribute to cardiac artifacts only.
