## Additional file 11 for "Solid-state esophageal pressure sensor for the estimation of pleural pressure: a bench and first-in-human validation study"

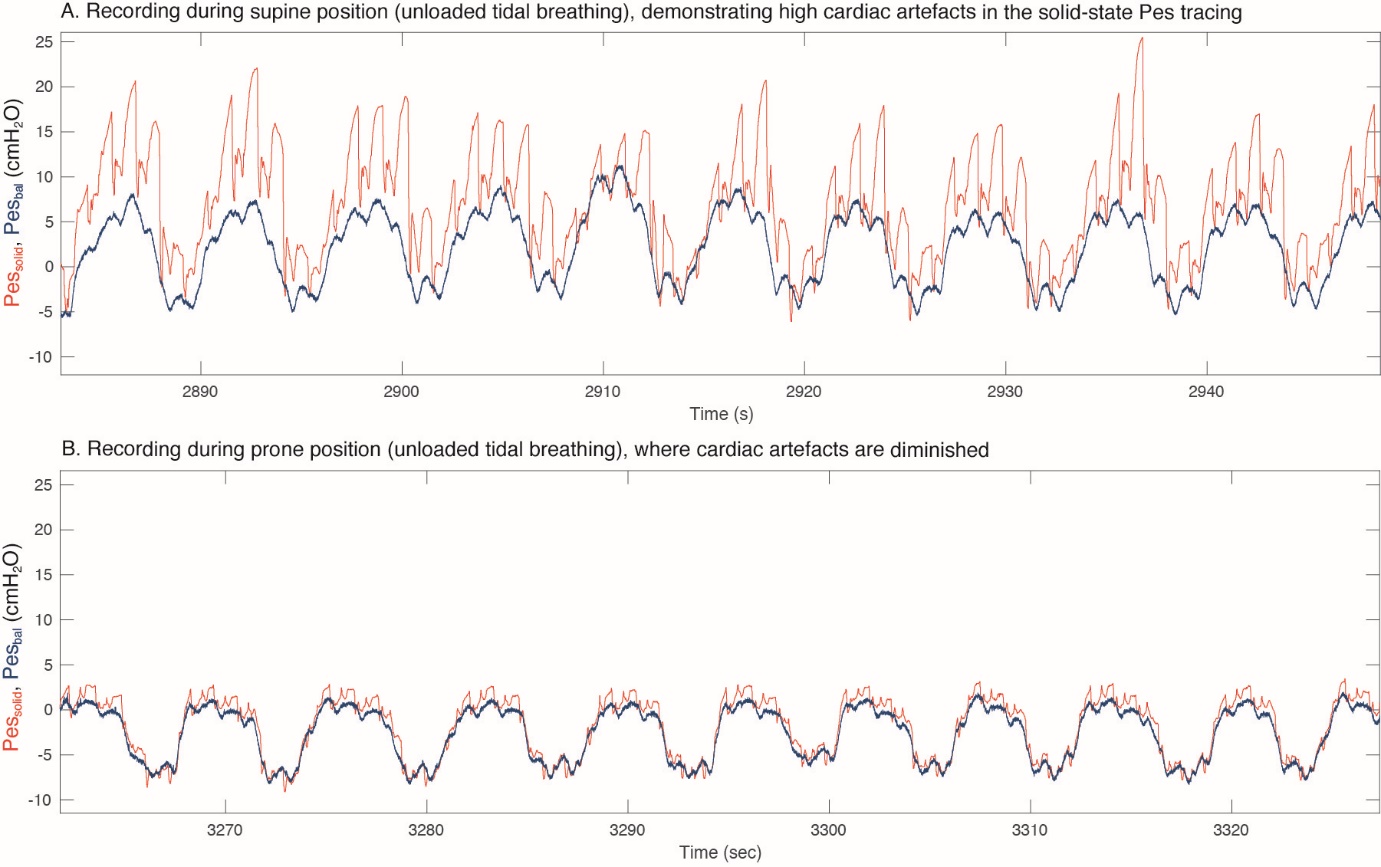

**Additional figure 11.** AB) Example of position-dependent artefacts in solid-state catheter
